# Estimation of SARS-CoV-2 antibody prevalence through integration of serology and incidence data

**DOI:** 10.1101/2021.03.27.21254471

**Authors:** Liangliang Wang, Joosung Min, Renny Doig, Lloyd T Elliott, Caroline Colijn

## Abstract

Serology tests for SARS-CoV-2 provide a paradigm for estimating the number of individuals who have had infection in the past (including cases that are not detected by routine testing, which has varied over the course of the pandemic and between jurisdictions). Classical statistical approaches to such estimation do not incorporate case counts over time, and may be inaccurate due to uncertainty about the sensitivity and specificity of the serology test. In this work, we provide a joint Bayesian model for case counts and serological data, integrating uncertainty through priors on the sensitivity and specificity. We also model the Phases of the pandemic with exponential growth and decay. This model improves upon maximum likelihood estimates by conditioning on more data, and by taking into account the epidemiological trajectory. We apply our model to the greater Vancouver area, British Columbia, Canada with data acquired during Phase 1 of the pandemic.

## 1 INTRODUCTION

SARS-CoV-2 has led to more than 2.5 million confirmed deaths globally and more than 114 million confirmed infections (Johns Hopkins University, 2020). As COVID-19 involves asymptomatic carriers and cases with mild symptoms (Day, 2020), infection of SARS-CoV-2 may be much more widespread than indicated by the number of confirmed cases. The aim of this work is to provide a Bayesian method for estimating the fraction of infections that were confirmed cases, i.e. the ascertainment fraction. We apply our method to serology and case count data from the greater Vancouver area, in British Columbia (B.C.), Canada collected during Phase 1 of the pandemic (the British Columbia Centre for Disease Control broadly divides the pandemic in B.C. into Phases covering periods with qualitatively similar epidemiological dynamics and measures). We combine serology measurements and case data using a Bayesian framework. An accurate estimate of ascertainment (and hence antibody prevalence) and serological accuracy could inform both policy and non-pharmaceutical interventions (NPI; Flaxman et al. 2020). Recent work in serology for COVID-19 has led to estimates of prevalence in Spain (Pollán et al., 2020) and New York (Stadlbauer et al., 2020). However, serology tests are imperfect. Without considering the accuracy of the test, conclusions based on serology measurements may be misleading. A recent study by the University of California in Santa Clara about serology measurements in Los Angeles has been criticized for their failure to incorporate accurate error rates in their results (Sood et al., 2020; McCormick, 2020).

Study of serology measurements must make reference to the sensitivity and specificity (the serological accuracy) of the test, incorporating uncertainty. We consider a Bayesian prior on these variable and also integrate data from confirmed cases. We provide a Bayesian model that incorporates these considerations and also incorporates an epidemiological model for infection dynamics. Our model is summarized in Figure 1. We compare our results to those in Skowronski et al. (2020). Our results are similar to what is found in Skowronski et al. (2020), but our confidence intervals are tighter.

**Figure 1:**
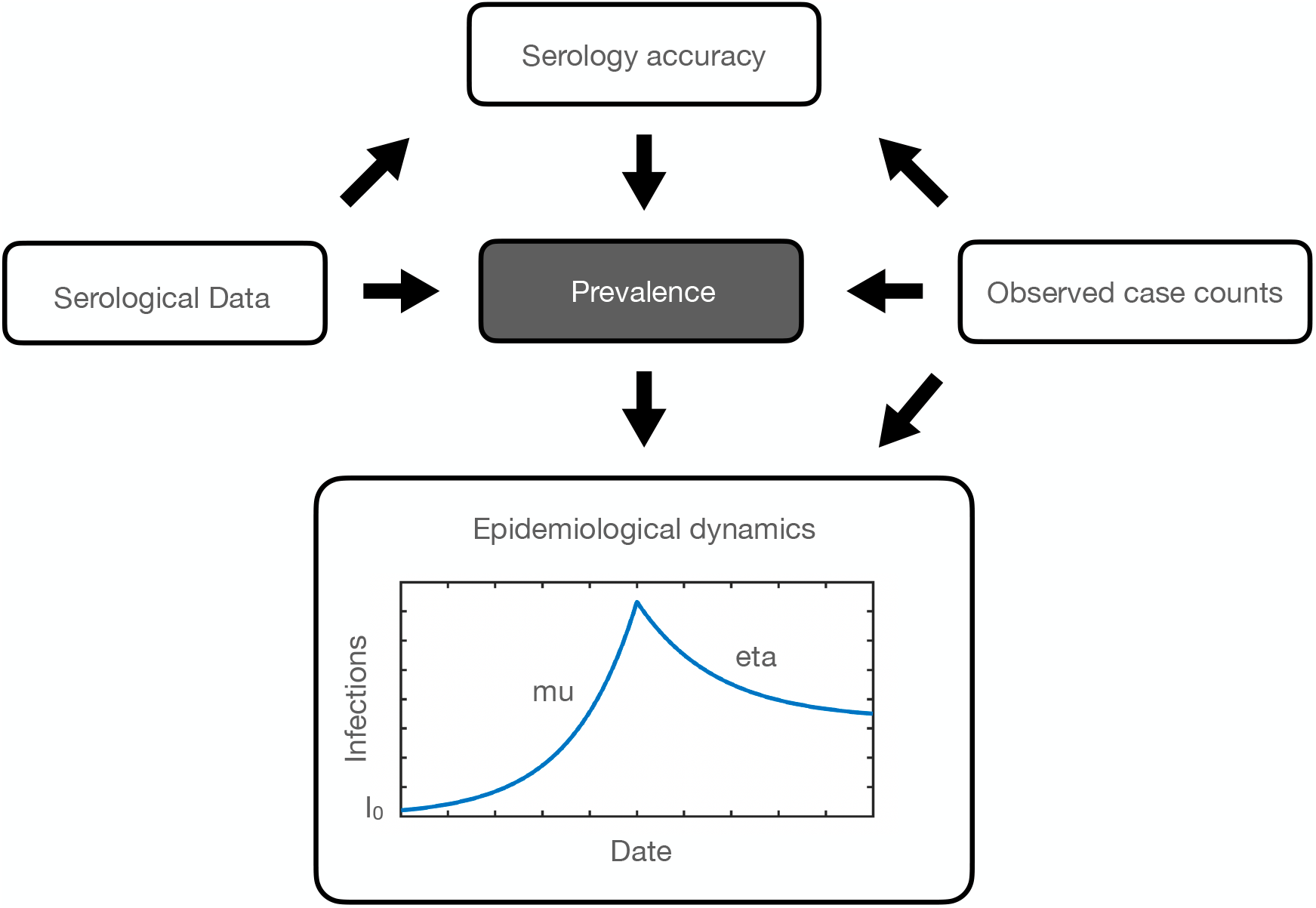
Plate diagram (Koller and Friedman, 2009) indicating the acyclic graph governing the relationships among variables in our model. Arrows indicate conditional dependencies, according to equations in the Methods Section. For epidemiological dynamics, we assume that the number of active cases increases exponentially with rate *µ* and then decreases according to exponential decay with rate *η* following the introduction of NPI measures. *I*_0_ is the number of initial cases among the population. This schematic represents antibody prevalence during a rise and then a fall of the pandemic over a single Phase. (For example, Phase 1 of the pandemic in B.C. from the end of January until the end of May.)

## 2 MATERIALS AND METHODS

Our model for antibody prevalence and the accuracy of serological assays is informed by two data modalities: confirmed cases of COVID-19 from testing, and serological data from serology surveys. In order to incorporate both types of data into our model, we define an underlying generative Bayesian process and then we derive Bayesian inference on the model parameters (including the antibody prevalence parameter).

We denote the number of confirmed cases at time *t* by *c*_*cc*_(*t*). These case counts are typically reported daily. To specify our generative process, we first define the process underlying the spread of active cases (the case count) in the population. We consider a model with exponential growth followed by exponential decay for the population dynamics for the number of cases. This model reflects the assumption that the reproduction number could be above one before sufficient NPI measures are instituted, and then below one after such measures are instituted.

Specifically, we use a 3-parameter exponential growth and decay model. We assume that at time *t* = 0 there are *I*_0_ infected individuals in the population. The number of infected individuals increases exponentially at rate *µ* > 0 until some time *τ*. After the time *t* = *τ*, the number of infected individuals is assumed to decrease exponentially with decay rate *η* > 0. This model is given by the following equation:

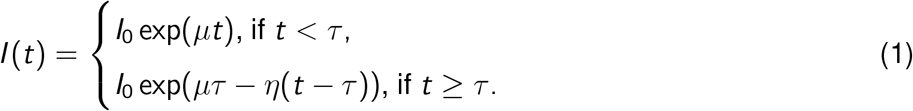

Here *I* (*t*) is the number of infected people at time *t*. We assume that some proportion, *p*_*cc*_, of these active cases are reported in the confirmed cases. Of interest for the modeling of serological data is the cumulative number of infections at time *t*. We define this generally as 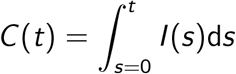. However in practice, since we have counts at discrete time points, this can be written as 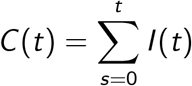.

Immunoassays are used to determine how many people have been infected with COVID-19, by testing if an individual has SARS-CoV-2 antibodies (we use data from Skowronski et al. 2020, which tests for S1 spike, nucleocapsid and S1 receptor binding domain by combining the Vitros XT 7600 analyzer, the ARCHITECT i2000SR analyzer from Abbot Laboratories and the ADVIA Centaur XPT system). This allows us to measure the sero-prevalence of SARS-CoV-2 antibodies in a given population at time *T*. These data consist of two integers: *m*_*p*_, the number of people who tested positive for SARS-CoV-2 antibodies and *n*, the total number of people tested. In order to analyse these results, we must take into consideration the accuracy of the testing procedure. To do this we consider the sensitivity and specificity of the immunoassays. The sensitivity of a test is the probability that an individual who has antibodies (D+) tests positive (T+; the true positive rate.) Specificity is the probability that an individual who has not contracted the illness (D-) test negative (T-; the true negative rate.) We denote each of these respectively as *S*_sens_ = *P*(*T* + |*D*+) and *S*_spec_ = *P*(*T* − |*D*−). When modeling the observed data, we must consider the overall probability that a given test comes back negative. The probability of a positive test can be expressed as:

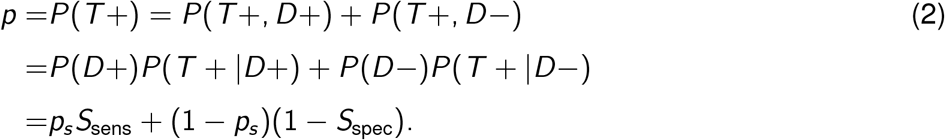

Here *p*_*s*_ is the probability that a randomly selected individual from the population has the antibodies. Relating the serological data to the cumulative confirmed cases can then be done by writing *p*_*s*_ = *C* (*T*)*/N*, where *N* is the total population from which the *n* serology tests were randomly administered.

### 2.1 A BAYESIAN MODEL FOR SEROLOGY

The model we propose here is a fully Bayesian hierarchical model for serological data (including an estimate of antibody prevalence at a single point in time). Our model is informed by epidemiology (in the sense of the known rise and fall). The likelihood for this model considers both confirmed cases and results from a serological survey. The first component of the likelihood arises from the case counts. These are assumed to follow a binomial distribution in which the probability of observation is *p*_cc_. We assume that there is some delay between the time when an individual is infected and the time when they are reported. Accordingly, the confirmed cases on a day *t* are distributed according to *c*_cc_(*t*) ∼ Binomial(*I* (*t* − *τ*_0_), *p*_cc_).

In the serological data, *m*_*p*_ is also assumed to follow a binomial distribution. Here the size parameter is the number of tests administered (*n*) and the binomial proportion is the probability of testing positive, *p*. These data are thus distributed according to *m*_*p*_ ∼ Binomial(*n, p*). Compressing notation, here we denote all observations as 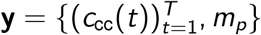 and all parameters as ***θ*** = {*µ, η, τ, τ*_0_, *I*_0_, *p*_cc_, *S*_sens_, *S*_spec_}. The full likelihood is thus:

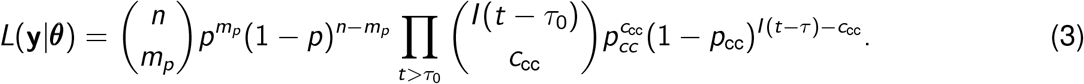

Here we have defined *p* in terms of **y** and ***θ***, as in Equation 2. Note from the above subsection that *p* depends on *C* (*T*), and *C* (*T*) depends on *c*_cc_, and so the likelihood in Equation 3 only splits conditionally on *C* (*T*) (indicating the conditional relationships displayed in Figure 1).

Three of our model parameters only take values in the (0, 1) interval: *p*_cc_, *S*_sens_, and *S*_spec_. For these parameters we assume a beta prior distribution with shape parameters 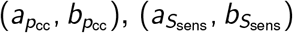, and 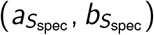 respectively. The remaining parameters can take values either in (0, ∞): (*µ* and *η*), or in [1, ∞): (*I*_0_, *τ*, and *τ*_0_). For these parameters we assign a normal distribution truncated appropriately on the left. Each of these parameters have hyperparameters for their means and standard deviations. Bayes’ rule can then be used to compute the posterior *p*(***θ***|**y**) from Equation 2 up to a constant of proportionality.

We perform posterior inference on this model using *Stan* (The Stan Development Team, 2020). *This is a probabilistic programming language that derives MCMC (Markov chain Monte Carlo) updates for estimating the posterior distribution of a generative process, conditioned on observations (Stan* is also used in other COVID-19 work such as Flaxman et al. 2020). Our code is available on *GitHub* under the open source BSD 2-clause license^1^.

## 3 EXPERIMENTS

We consider simulation studies, and an experiment about prevalence and serological accuracy in the Greater Vancouver Area in B.C., Canada.

### 3.1 SIMULATION STUDIES

To assess the identifiability of our model, we simulate data according to the parameter settings displayed in Table 1 (these are the same parameters that we use for our experiment on real B.C. serological data). We simulate confirmed cases along a trajectory of 120 days (*t* = 1, …, 120). From the parameter settings in Table 1 we can compute the trajectory of infections *I* (*t*) through Equation 1. The sero-prevalence is computed as 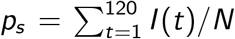 (where *N* is the population size) and the probability of a positive serological test is computed from the relationship defined in Equation 2. We then generate a sequence of case counts and the number of positive serology tests from the binomial distributions as defined in Section 2.1. We simulate 10 datasets using the settings in Table 1, and aggregate our results over these 10 independent datasets.

**Table 1:**
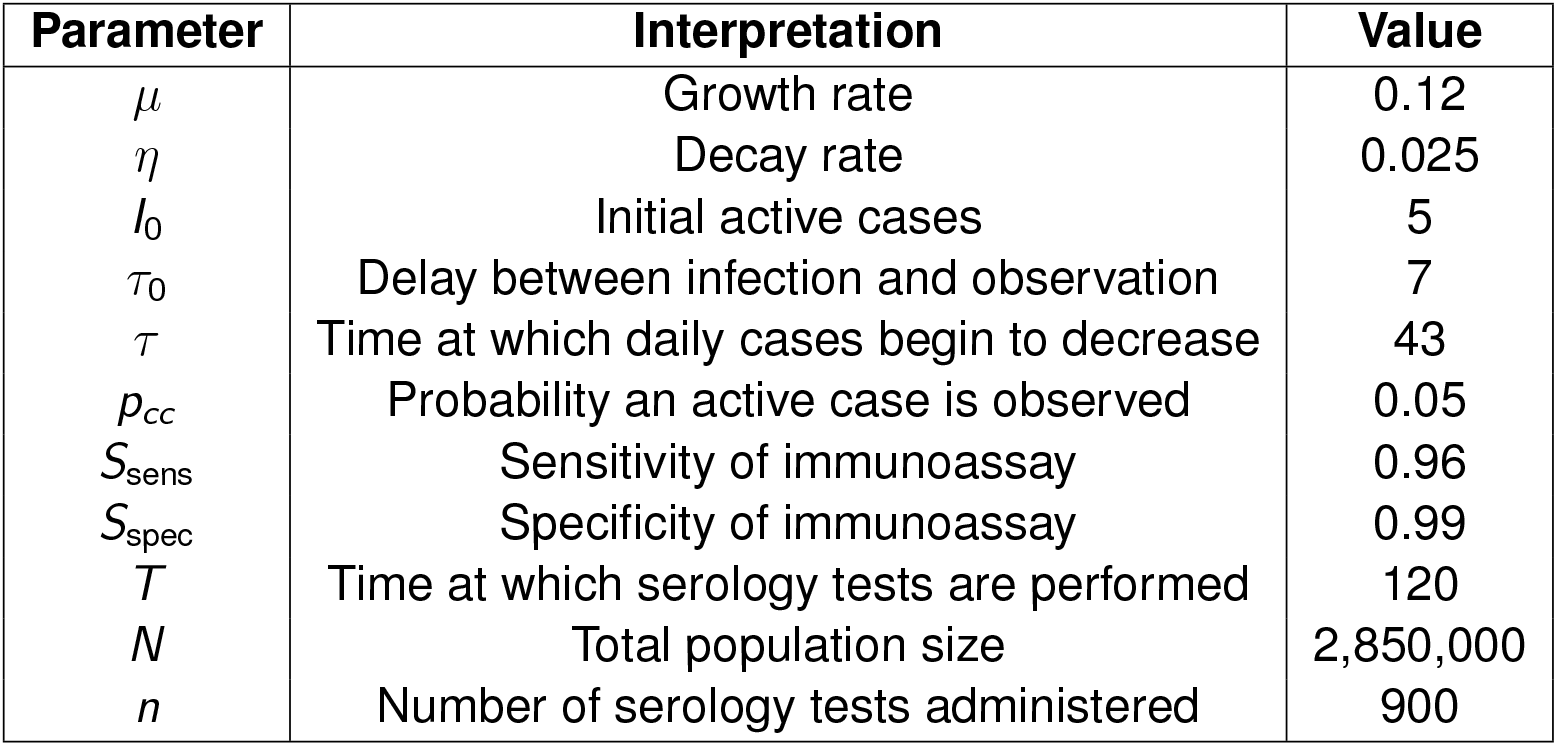
Parameter settings for simulation study, based on priors used for the B.C. data.

| Parameter | Interpretation | Value |
| --- | --- | --- |
| $\mu$ | Growth rate | 0.12 |
| $\eta$ | Decay rate | 0.025 |
| $I_0$ | Initial active cases | 5 |
| $\tau_0$ | Delay between infection and observation | 7 |
| $\tau$ | Time at which daily cases begin to decrease | 43 |
| $p_{cc}$ | Probability an active case is observed | 0.05 |
| $S_{sens}$ | Sensitivity of immunoassay | 0.96 |
| $S_{spec}$ | Specificity of immunoassay | 0.99 |
| $T$ | Time at which serology tests are performed | 120 |
| $N$ | Total population size | 2,850,000 |
| $n$ | Number of serology tests administered | 900 |

Through this simulation study, we aim to answer two main questions: 1) How accurately do the posterior means estimate the true parameter values, and how well does the 95% credible interval capture the true values? 2) To what extent does the choice of prior distribution affect the results? To this end, we fit our model under four different prior settings. We first distinguish the parameters into “positive-valued”, *µ, η, I*_0_, *τ*_0_, *τ*, and “0-1-valued”, *p*_cc_, *S*_sens_, *S*_spec_. For the positive-valued parameters we compare truncated normal, gamma, and exponential priors, while keeping the 0-1-valued priors set according to a beta distribution. Similarly, for the 0-1-valued parameters we compare truncated normal and beta priors, while keeping the positive-valued priors set to a truncated normal. We select the prior parameters in such a way that the prior mean and variance is matched across all prior settings (matching Table 1). The only exception to this matching is the exponential prior, whose variance depends strictly on the mean and thus cannot be matched.

Simulation results for each prior setting are summarized in Figure 2. According to that Figure, the 95% confidence intervals for the posteriors in these simulations overlap with the true parameter values in all cases. This suggests that our methods are not sensitive to the priors chosen. A grid of pairwise scatterplots of the posterior samples for *I*_0_, *p*_cc1_, *p*_cc2_, and *µ* is shown in Figure 3. These scatterplots can be used to evaluate the relationships between the parameters of our model. The figure shows that *p*_*cc*1_ and *p*_*cc*2_ are highly correlated. There also appears to be some nearly linear relationship between *I*_0_ and *µ*. This suggests that there could be identifiability issues between these parameters.

**Figure 2:**
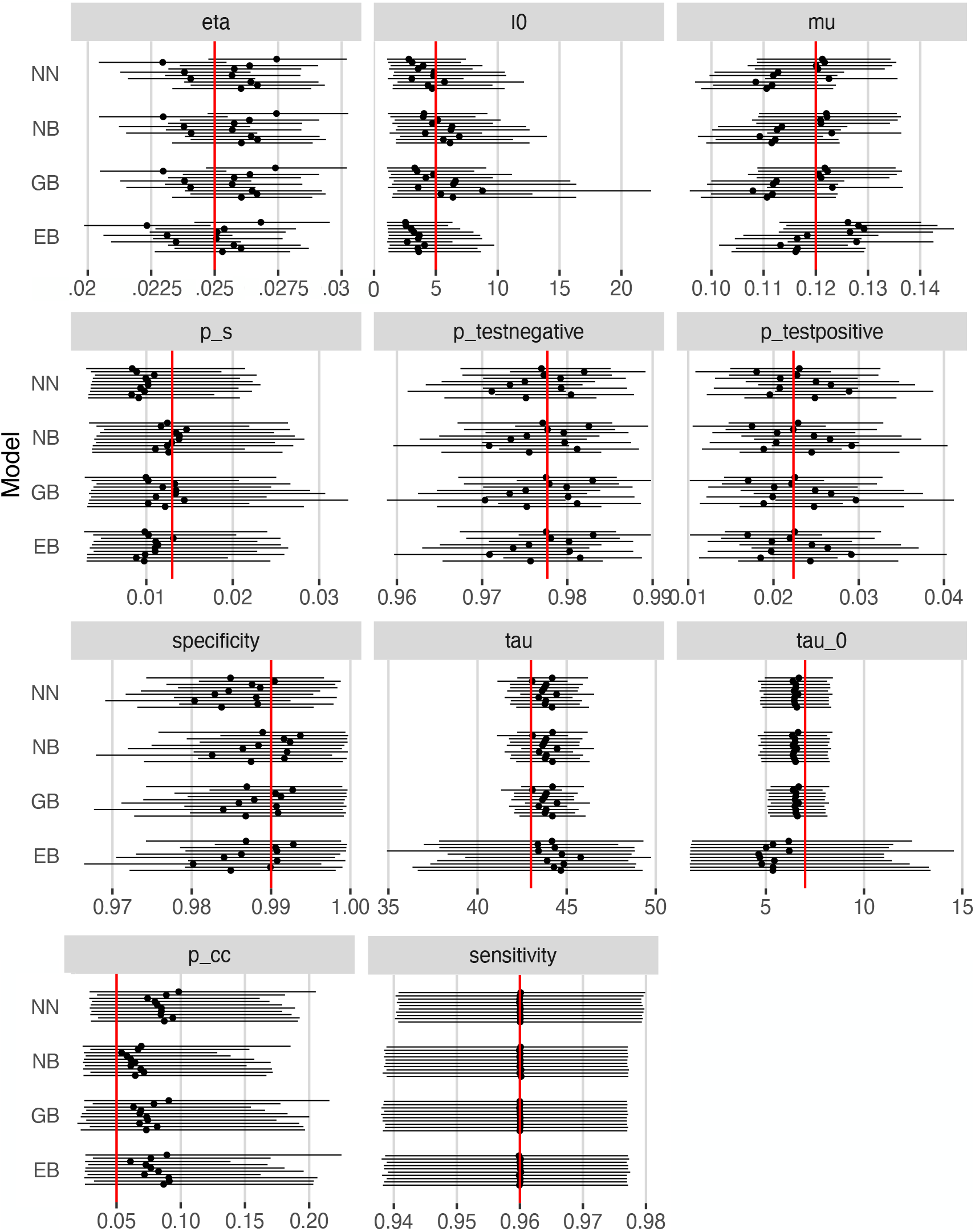
95% credible bands (solid lines) and posterior means (solid points) under each prior setting. Priors used were truncated normal/beta (NB), gamma/beta (GB), exponential/beta (EB), and truncated normal/truncated normal (NN) for the positive-valued/0-1-valued prior distributions respectively. Red lines indicate true parameter values.

**Figure 3:**
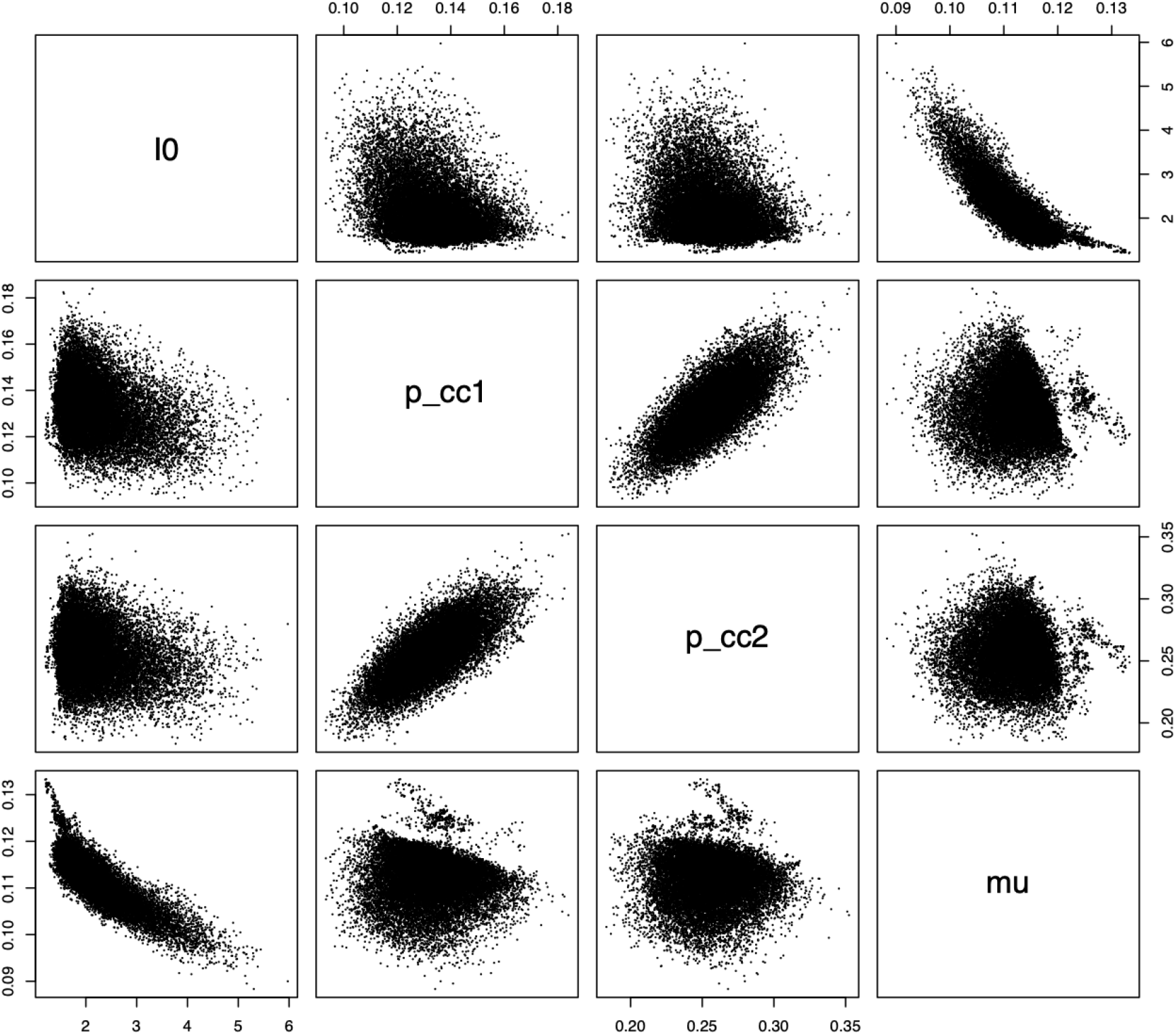
Pairwise scatterplots of the posterior samples from our simulation study. A beta prior distribution was used for *p*_*cc*1_ and *p*_*cc*2_, and a truncated normal prior was used for *I*_0_ and *µ*. Strong correlation is indicated between *p*_cc1_ and *p*_cc2_.

### 3.2 B.C. PREVALENCE AND ACCURACY FROM SEROLOGY

We analyze data from the greater Vancouver area in B.C., Canada for Fraser and Vancouver Coastal Health Authorities. This analysis is based on serological data from Summer 2020 acquired from Skowronski et al. (2020). We use confirmed case data from the BCCDC (British Columbia Centre for Disease Control, 2020). This daily confirmed case count data starts from January 26, 2020. The Skowronski et al. (2020) study shows that in the greater Vancouver area, out of 885 people tested between May 15 and May 27, four tested positive. For this analysis we assume the population of the greater Vancouver area around that time is 2.85 million (The Government of British Columbia, 2020). To determine settings for the exponential decay rates and growth rates, we referred to simulations we performed for which *µ* = 0.1 and *η* = 0.05 (these are the growth rate and decay rate for a Phase of the pandemic in B.C.). These growth and decay rates are sourced from Anderson et al. (2020). To account for the uncertain nature of the prior data we assign a standard deviation of 0.1 for the parameters *µ* and *τ*. The mean values for these estimates are also obtained from Anderson et al. (2020) and references therein. In that work, the estimate of the delay between infection and reporting was found to be approximately one week. Cases in B.C. started to decrease around mid to late March; giving us a reasonable mean prior value for *τ* at 50 days since January 26, 2020. We set the prior mean for *I*_0_ (the initial number of infections) to reflect the estimate of 8 active cases in B.C., as that number was reported on February 1st, 2020 (Anderson et al., 2020).

In B.C., testing protocols were changed on April 14 to increase the testing rate (British Columbia Centre for Disease Control, 2020). For this reason we modify the likelihood to incorporate two testing rates, *p*_cc1_ and *p*_cc2_ (the testing probability that a case is observed through a polymerase chain reaction, or PCR, test before and after April 14). Computing the binomial likelihood for the date *t* prior to April 14 will use *p*_cc1_ and after that date we will use *p*_cc2_. The prior hyperparameters for these variables are set to (35, 65) and (65, 35) respectively, yielding a mean of 0.35 for *p*_cc1_ and 0.65 for *p*_cc2_. These two values are found as the estimated sampling proportions for B.C. in Anderson et al. (2020). While our model is different from the model of Anderson et al. (2020), the interpretation of our parameters is similar. Based on the sensitivies and specificities reported in Skowronski et al. (2020), we selected prior parameters that yielded a mean of 0.9 and 0.99 respectively. The variance for both parameters was set to be 0.009; the corresponding shape parameters were computed from the mean and variance.

## 4 RESULTS

In Figure 4, we provide our estimate of the number of active cases over the time period examined, according to our posterior samples. Our results indicate that the prevalence of SARS-CoV-2 antibodies during Phase 1 of the pandemic in B.C. indicates that approximately 0.57% of the population was infected (95% confidence interval [0.48%, 0.68%]). In Figure 5 we provide histograms of the posterior samples from all of the model parameters. This includes the parameters for which we have defined priors, as well as *p*_*s*_, *p*_testnegative_, and *p*_testpositive_. The 95% credible intervals for the posteriors for all of the parameters are provided in Table 2.

**Table 2:** Posterior mean, median, and 95% credible interval for model parameters and derived parameters for B.C. serological data. *µ* and *η* indicate the exponential rise and fall of cases. *I*_0_ indicates the number of cases at the first time point. *τ* indicates the time at which the number of cases changes from exponential growth to exponential decay. *τ*_0_ indicates the number of days between infection of an individual and reporting. *p*_cc1_ and *p*_cc2_ indicate the probability of observation (before and after testing procedures were changed). *S*_sens_ and *S*_spec_indicate the sensitivity and specificity of the serological assay. *p*_*s*_ indicates the sero-prevalence.

| Parameter | Mean | Median | 95% Credible Interval |
| --- | --- | --- | --- |
| $\mu$ | 0.104 | 0.104 | (0.095, 0.118) |
| $\eta$ | 0.052 | 0.052 | (0.048, 0.057) |
| $I_0$ | 4.612 | 4.620 | (2.445, 6.741) |
| $\tau$ | 46.517 | 46.519 | (45.029, 47.988) |
| $\tau_0$ | 5.111 | 5.117 | (3.694, 6.514) |
| $p_{cc1}$ | 0.107 | 0.107 | (0.088, 0.128) |
| $p_{cc2}$ | 0.339 | 0.339 | (0.286, 0.395) |
| $S_{sens}$ | 0.898 | 0.924 | (0.655, 0.998) |
| $S_{spec}$ | 1.000 | 1.000 | (0.999, 1.000) |
| $p_s$ | 0.006 | 0.006 | (0.005, 0.007) |
| $p_{testnegative}$ | 0.995 | 0.995 | (0.993, 0.996) |
| $p_{testpositive}$ | 0.005 | 0.005 | (0.004, 0.007) |

**Figure 4:**
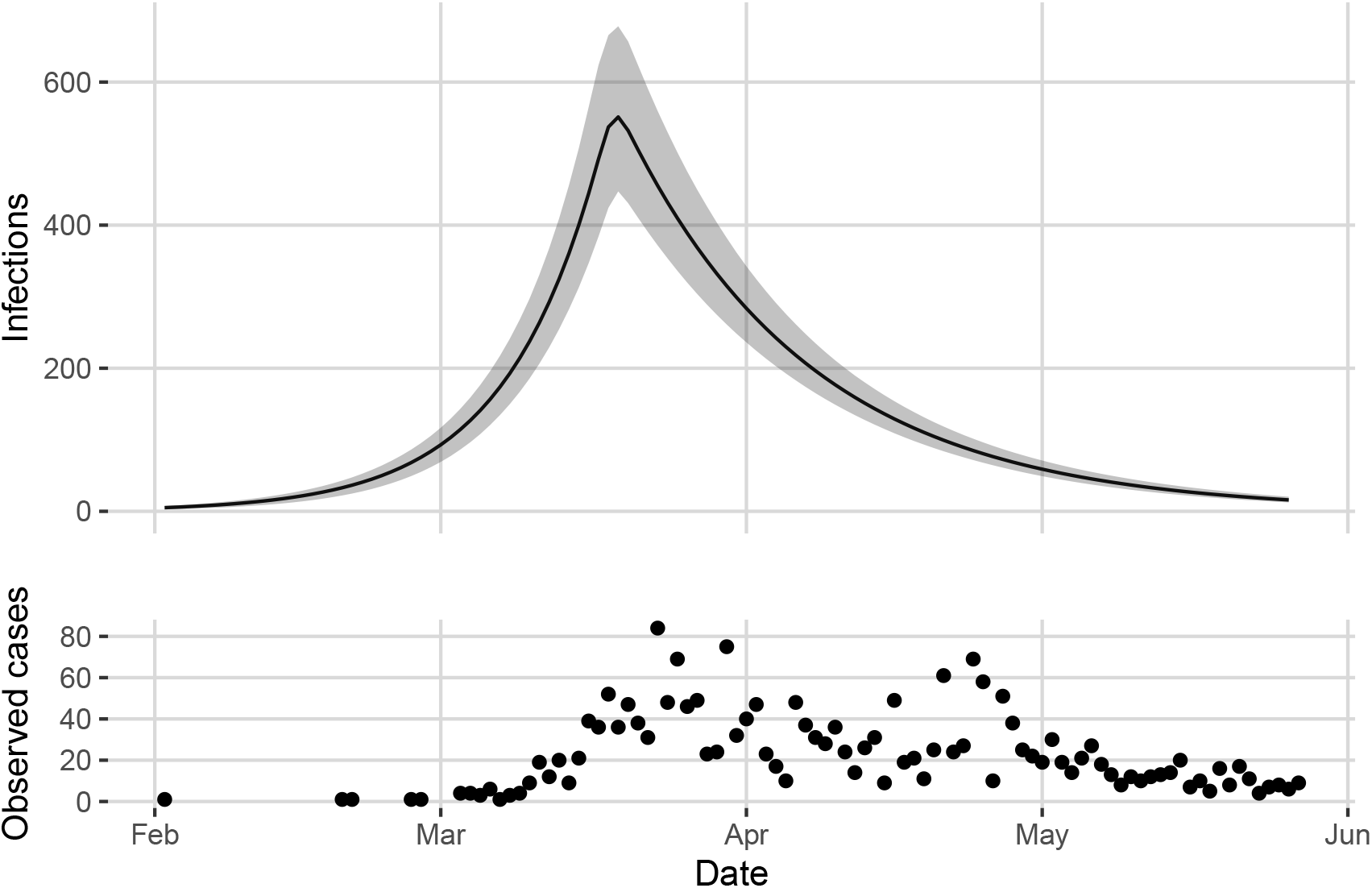
Top: Estimated number of infections in the greater Vancouver area between January 26 and May 27. Posterior mean shown by the solid line with 95% credible band in grey. Bottom: Observed cases in Vancouver over the same time frame.

**Figure 5:**
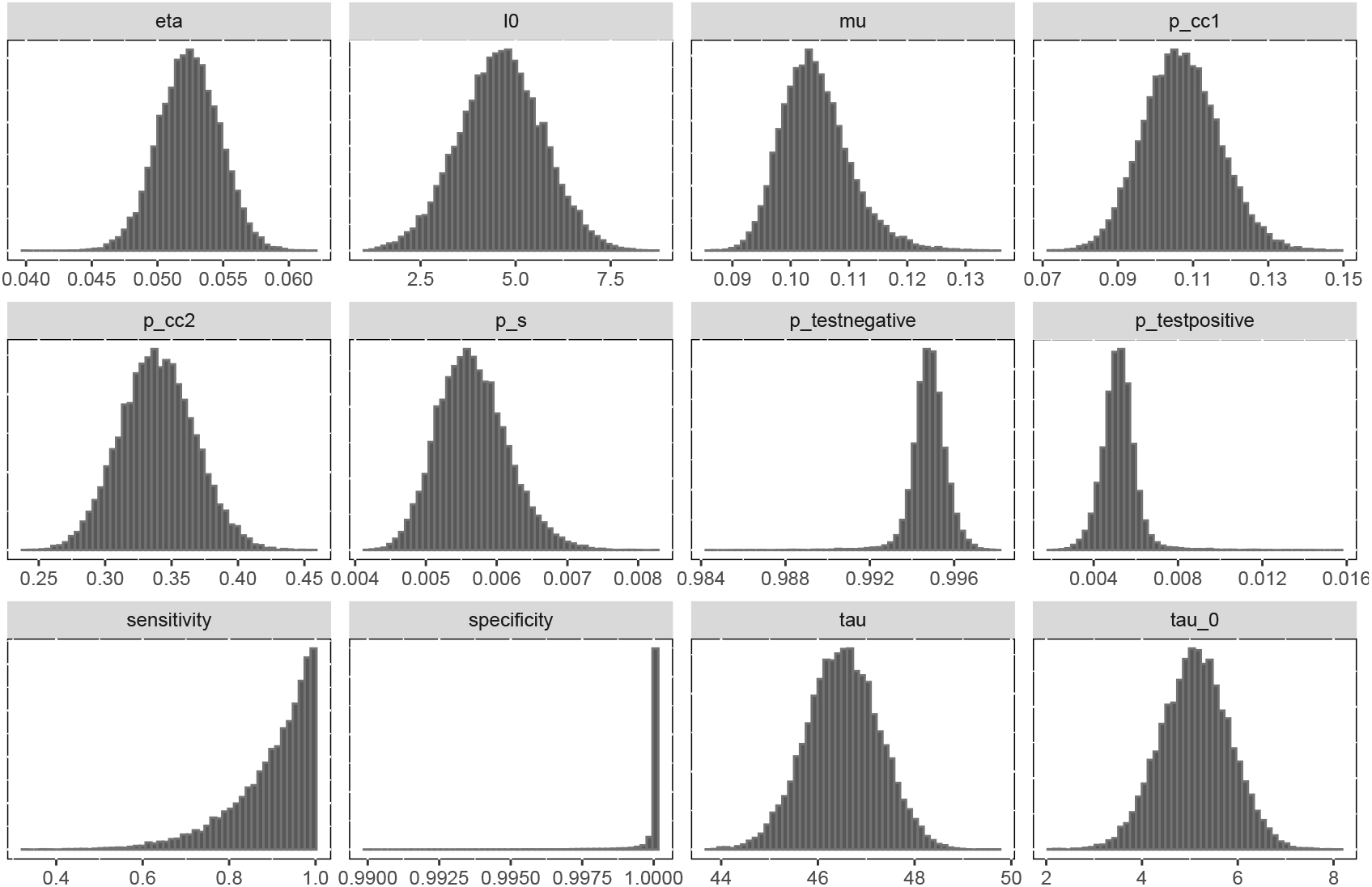
Posterior samples for all model parameters plus derived parameters *p*_*s*_, *p*_testnegative_, and *p*_testpositive_ for the experiment on B.C. serological data. All parameters have unimodal distributions, and there is evidence of MCMC mixing.

From the model parameters we can obtain samples from the posterior distribution of the number of active cases between January 26 and May 27. The posterior mean and 95% credible interval are shown in Figure 4 along with the observed case counts.

Table 3 summarizes the posterior distribution of the probability of case detection before and after April 14 (*p*_*cc*1_ and *p*_*cc*2_), as well as the cumulative number of cases in the greater Vancouver area between January 26 and May 27. The cumulative number of cases (up to and including May 27) reflects the number of daily cases from our model resulting from the estimated model parameters. We find that the maximum number of simultaneous infections (including unobserved cases) during this Phase occurred on March 19th (552 infections, 95% confidence interval [446, 678]).

**Table 3:** Posterior mean, median, and 95% credible interval for *p*_*cc*1_, *p*_*cc*2_, and *C* (the cumulative count on May 27) for Greater Vancouver Area serological data.

| Parameter | Mean | Median | 95% Credible Interval |
| --- | --- | --- | --- |
| $p_{cc1}$ | 0.107 | 0.107 | (0.088, 0.128) |
| $p_{cc2}$ | 0.339 | 0.339 | (0.286, 0.395) |
| $C$ | 16,150 | 16,060 | (13,651, 19,193) |

These results also suggest that on May 27 (the last day of the Phase we investigate) the posterior mean percentage of people in the greater Vancouver area who had been infected with COVID-19 was around 0.57% of the total population, approximately eight times the reported number of infections over the same time period. Previously, Skowronski et al. (2020) estimated a sero-prevalence of 0.55% in the greater Vancouver area at end of May 2020 (using conventional methods, without a connection to confirmed case counts from polymerase chain reaction tests or disease dynamics). These results show that principled and advanced Bayesian methods incorporating case counts and disease dynamics broadly match the conventional methods for estimation of sero-prevalence reported in Skowronski et al. (2020).

## 5 DISCUSSION AND CONCLUSION

In this work, we have presented a Bayesian model for serological data, integrating test sensitivity and specificity with an epidemiological model for case counts. Our model improves upon previous work in serology measurements (Sood et al., 2020; McCormick, 2020) by incorporating uncertainty about testing accuracy. Bayesian methods allow higher accuracy than maximum likelihood methods in posterior estimation of parameters, through integration over uncertainty. Our prior includes an epidemiological model involving an increase followed by a decrease in case counts. This approximation (an increase, followed by a decrease) is consistent with the Phases of COVID-19 case counts in B.C. and in other locations; our work involves examination of COVID-19 data restricted to a single Phase.

In B.C. testing policy was modified mid April to widen testing to anyone with COVID-19-specific symptoms, overlapping with the Phase we consider. We model this change by allowing the probability that an active case is observed to vary on either side of the change in testing procedures. Testing procedures are otherwise broadly constant (and focused on PCR testing of individuals with symptoms of COVID-19). Our procedure mitigates inaccuracies in estimation that could arise if the testing fraction were assumed to be constant.

Our methods could easily be extended to model multiple Phases by coupling the parameters from one Phase to the next (with change points constructed at dates reflecting inflection points identified by external model fits or by the times at which policies were changed). Alternatively, change points could be added as free variables in the *Stan* estimation code (The Stan Development Team, 2020), including the rise and fall of case counts independently over each learned range. In addition, the models we propose could be stratified by age, or extended to include hospitalization, ICU (intensive care unit) data and deaths, through a more complex Bayesian model over these multiple modalities.

Our methods allow serological data to be combined with COVID-19 case counts from PCR tests, to better estimate prevalence of an antibody response, to estimate the fraction of infections that were detected, and to extrapolate future trends in the pandemic. We report a posterior mean antibody prevalence of 0.57% in the greater Vancouver area during Phase 1 of the pandemic, which broadly matches previous report of 0.55% derived from the same serological data (Skowronski et al., 2020). The interval reported in Skowronski et al. (2020) was [0.15%, 1.37%]. Our confidence interval is tighter (varying by 0.19% compared to 1.22% for Skowronski et al. 2020), and nested inside the confidence interval reported in Skowronski et al. (2020). Our posterior conditions on more data (case counts) and is constrained by disease dynamics, and so we expect less uncertainty in our estimates (this is reflected in our tighter confidence intervals). For our experiment on the Greater Vancouver area, our posterior mean matches classical methods (Skowronski et al., 2020), but our estimate is more precise.

## Data Availability

Open source software reproducing our experiments are provided on GitHub.

https://github.com/lell/serology

## Footnotes

1 Available at https://www.github.com/lell/serology/

